# A novel class of non-coding variants driving DNA double-strand breaks is associated with complex genetic diseases

**DOI:** 10.1101/2025.10.16.25338147

**Authors:** Sébastien Auber, Sarah Collins, Andrey Buyan, Ariuna Aiusheeva, Anne-Laure Finoux, Florian Saur, Sarah Cohen, Vincent Rocher, Coline Arnould, Marie Verbanck, Ivan V. Kulakovskiy, Gaëlle Legube, Raphaël Mourad

## Abstract

Previous research efforts to map the genetic determinants of complex diseases genome-wide have identified genetic variants that act at the protein level by disrupting function, or at the regulatory level by affecting gene expression. However, for most mutations, the underlying molecular mechanisms remain unknown. In addition to protein function and gene expression level, complex diseases have previously been associated with DNA double-strand breaks (DSBs), particularly in the case of cancer, immune disorders, and neurological and psychiatric diseases. Such DSBs have the potential to regulate or interfere with transcription, replication, and genome maintenance. However, no mutations that alter the occurrence of DSBs have yet been reported. By mapping genome-wide SNPs that alter the binding of DNA repair proteins, we discovered a new class of non-coding SNPs that we named dsbSNPs and which modulate DSB frequency. Importantly, while some DSBs occur in transcribed chromatin, transcription does not seem to have a causal effect on DSBs. Instead, dsbSNPs can influence the selection of the repair pathway depending on whether they alter transcription factor binding or not, thereby potentially controlling the mutation rate in the vicinity of DSBs. dsbSNPs showed high enrichment for GWAS top associations, twice as high as for eSNPs. Such associations support a novel mechanism by which non-coding variants altering DSB formation may play a role in complex genetic diseases.

## Introduction

Double-strand breaks (DSBs) occur when both DNA strands of the double helix are cleaved. DSBs can occur due to exogenous sources (*e.g.*, chemotherapy or radiation) or endogenous sources such as replication fork collapse or incomplete topoisomerase reaction. In recent years, genome-wide mapping of DSBs, such as BLESS, BLISS, and END-seq, has identified tens of thousands of endogenous DSB hotspots in humans, which correspond to DSBs generated under physiological conditions^1–3^. DSBs have been associated with genomic instability^4^ and are therefore particularly detrimental to the cell.

Genome instability is a hallmark of numerous complex genetic diseases, including certain neurodegenerative and psychiatric disorders, (auto)-immune disorders, and cancer^5–8^. Among these genetic diseases, genome instability has often been attributed to inherited mutations in DSB repair genes, such as BRCA1, BRCA2, ATM, 53BP1, Lig4, MDC1, and NBN^9,10^. In addition, over the past decades, genome-wide association studies (GWASs) have successfully identified hundreds of thousands of associations between complex diseases and single nucleotide polymorphisms (SNPs) located in numerous loci, often but far from exclusively mapping to some DNA repair genes. Importantly, more than 95% of these SNPs are located in non-coding sequences, making the elucidation of the underlying biological mechanism a major challenge^11,12^. Indeed, a systematic review identified only 309 experimentally validated non-coding regulatory GWAS SNPs^13^. Recent large-scale studies by consortia such as Genotype-Tissue Expression (GTEx) have shown that SNPs affect gene expression in many tissues, potentially contributing to disease susceptibility^14^. Other large-scale studies reported that SNPs could affect histone modification, RNA polymerase II binding, transcription factor binding or splicing^15–18^. However, recent attempts to map GWAS SNPs to functional SNPs have been underwhelming, showing a lack of colocalization between GWAS and eSNPs (expression SNPs)^19^ and even systematic structural differences^20^. Similarly, a systematic comparison of GWASs and EWASs (epigenome-wide association studies) showed they do not identify the same causal genes^21^. Moreover, a major additional challenge that prevents pinpointing causal variants from GWASs is linkage disequilibrium (LD), which corresponds to the non-random association of alleles of neighbouring SNPs. LD greatly complicates the interpretation of GWAS results, making candidate causal SNPs in a genomic locus indistinguishable. To prioritize causal SNPs within a locus, the most prominent method is statistical fine-mapping^22^; however, it heavily relies on genomic annotations.

On a different note, previous studies mapped variants associated with replication timing. Given that replication timing was associated with locus-specific mutation rates, the authors hypothesized that replication timing variants have the potential to affect mutation rates in their vicinity^23,24^. However, they could not show that DNA variants can affect DSB occurrence, which, in turn, may influence mutation rates. And no attempt to link such variants to genetic diseases was undertaken.

In a previous *in silico* study, we demonstrated that DNA motifs, including those of CTCF and several transcription factors, are predictors of endogenous DSBs^25^. Another computational study has shown the association of predicted non-canonical secondary structures (non-B DNA, including G4) with increased mutagenesis^26^. These studies suggest that DNA variants that affect CTCF binding, transcription, or non-B DNA structures may have a positive or negative impact on DSB frequency and, consequently, on diseases associated with genome instability-such as cancer.

Here, we have comprehensively demonstrated the link between DNA sequence and endogenous DSBs using *in silico* experiments based on deep learning models trained on DSB mapping datasets. Using SNP allelic imbalance from a set of ChIP-seq experiments for DSB repair proteins, we further report the existence of a novel class of non-coding SNPs involved in DSB frequency, which we further call dsbSNPs. Such dsbSNPs are located in regulatory elements (especially active promoters) and heterochromatin regions. Motif analysis at dsbSNPs did not reveal a causal role of transcription and R-loops on DSB formation, but instead strongly suggested that dsbSNPs affect DSB the selection of the repair pathway. Finally, we report the association of dsbSNPs with many complex diseases, suggesting a central role of dsbSNPs in their etiology.

## Results

### An additional paradigm: The dsbSNP hypothesis

We illustrated DSB recognition and repair, and the key proteins involved in Fig. 1a^27,28^. Following DSB formation, ATM is phosphorylated (pATM) and binds to the DSB. Then, two distinct pathways can repair the DSB: the non-homologous end-joining (NHEJ) and the homologous recombination (HR) pathways. In mammalian cells, NHEJ is favored over HR and occurs in all phases of the cell cycle. NHEJ involves factors such as KU70/KU80 that rapidly recognize broken DNA ends and protect them from nuclease activity. Next, XLF interacts with the XRCC4/Lig4 complex for ligation. 53BP1 plays a key role in promoting NHEJ by protecting DNA ends from resection, thus antagonizing HR. Conversely, HR is mostly active during the S and G2 phases. MRN recognizes DSBs, and resection takes place. BRCA1 promotes end resection and facilitates the recruitment of repair proteins, including BRCA2. The RPA protein (replication protein A) binds to single-stranded DNA, and then mediator proteins like BRCA2 favor RPA displacement by RAD51 to initiate strand invasion in the sister chromatid. In addition, SETX resolves R-loops, which are RNA-DNA hybrid structures that can form during transcription and interfere with DNA repair^29,30^.

**Figure 1:**
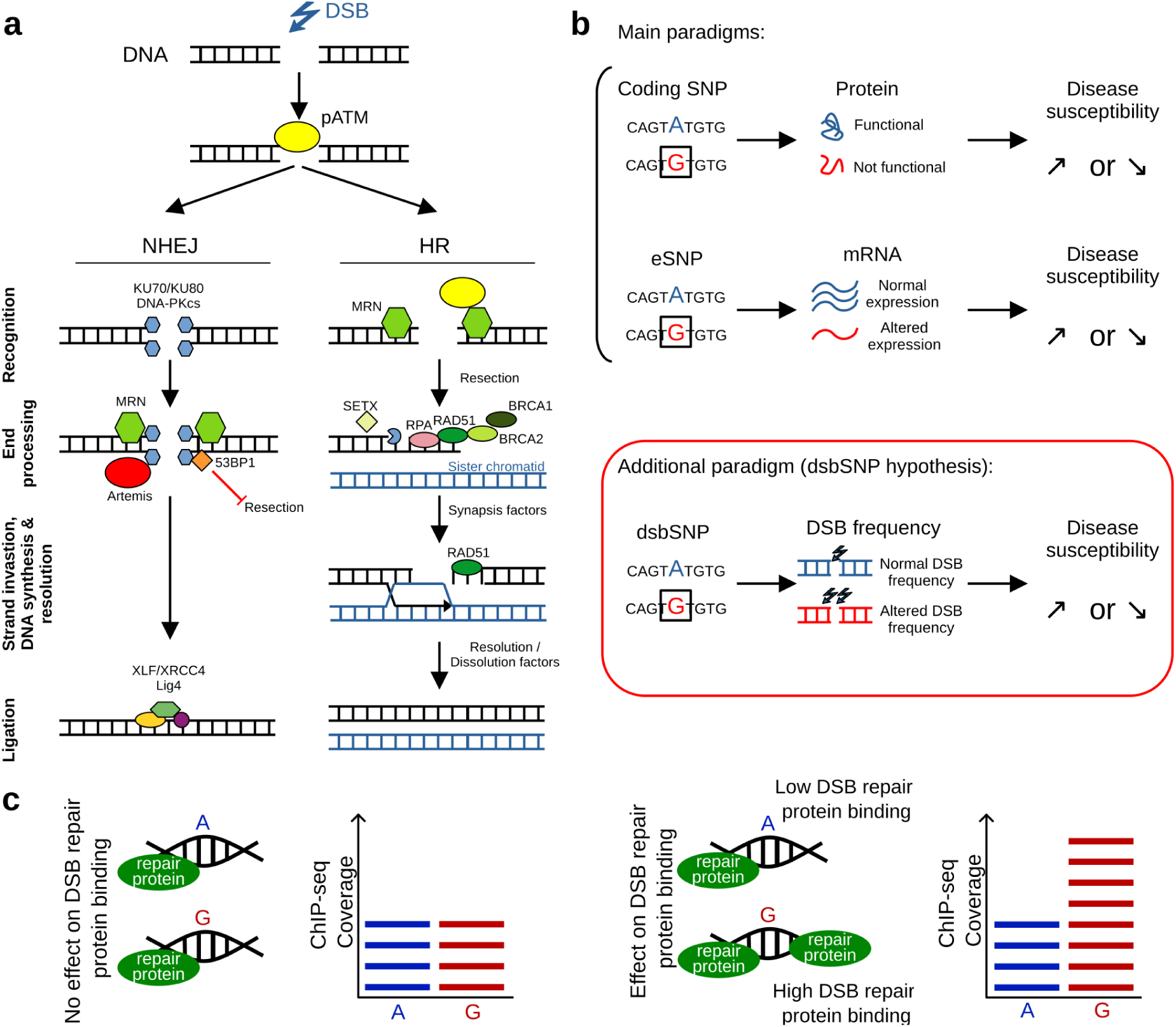
The dsbSNP hypothesis and DSB repair factors. **a)** Representation of the two main DSB repair pathways: Non-Homologous End-Joining (NHEJ) and Homologous Recombination (HR). **b)** The dsbSNP hypothesis in the context of complex genetic diseases. **c)** Illustration of the detection of allelic imbalance for a SNP using a DNA repair binding protein ChIP-seq.

In complex genetic diseases, there are two main paradigms explaining the association of variants, such as SNPs (Fig. 1b): (i) a SNP located in a coding sequence (or outside if it affects splicing) alters the protein sequence, which increases or decreases disease susceptibility; (ii) a SNP located in a non-coding sequence modifies the expression of a gene (*e.g.*, the mRNA level), which then affects disease susceptibility. Here, we propose an additional paradigm: we hypothesize the existence of a novel class of non-coding SNPs involved in DSB frequency, the so-called dsbSNPs. In this paradigm, a non-coding SNP can alter DSB frequency, which further affects disease susceptibility.

The putative dsbSNPs can be detected by estimating the allelic imbalance between homologous chromosomes: with large-scale ChIP-Seq data, the detection of significant differences in DSB repair protein binding between the two alleles (reference and alternative alleles) at heterozygous loci within a single genome is possible (Fig. 1c). The effect size (ES) measures how much the alternative allele is associated with increased (ES>0) or decreased (ES<0) protein binding, as compared to the reference allele. Importantly, our approach is performed at a very high resolution and allows us to precisely pinpoint dsbSNPs within ChIP-seq peaks of ∼250 bp.

### DNA sequence contributes to endogenous DSB frequency

We first comprehensively assessed the importance of DNA sequence as a determinant of endogenous DSB hotspots using recent deep learning models^31^ and diverse genome-wide DSB mapping techniques (listed in Materials and Methods, DSB mapping). On the one hand, we used DSBCapture, which maps DSBs independently of DNA repair proteins (in NHEK cells). On the other hand, we used ChIP-seq of DNA repair proteins: pATM and 53BP1 (in U2OS-DIvA cells), BRCA1 and NBN (in GM12878 cells), and BRCA2 (in hTERT-HME1 cells) (see Section Materials and Methods). Of note, the U2OS-DIvA cells, which were originally developed to induce sequence-specific DSB upon hydroxytamoxifen addition^32^, were not treated with hydroxytamoxifen here. Using the different DSB mapping data, we trained DeepSea models^33^ to predict the presence or absence of DSB mapping peaks from the DNA sequence. Here, each model was trained and tested on the same protein and cell line.

DSBCapture peaks could be predicted by the underlying DNA sequence (AUROC=0.86; AUPR=0.41; Fig. 2a, left side, and S1a). Interestingly, we observed a stronger prediction accuracy using pATM ChIP-seq data (AUROC=0.97; AUPR=0.28). We further explored other DSB repair proteins individually, including BRCA1, BRCA2, 53BP1, NBN, and again found good prediction accuracies. As an example, when looking at *MYC* and *IFNAR1* loci, we found that the two strongest endogenous DSB hotspots were accurately predicted (Fig. 2b, in red). We then used the model trained on pATM ChIP-seq in U2OS-DIvA cells and observed that it could predict not only pATM signals, but also 53BP1, END-seq, and BLESS signals in the same cells (Fig. 2c, left).

**Figure 2:**
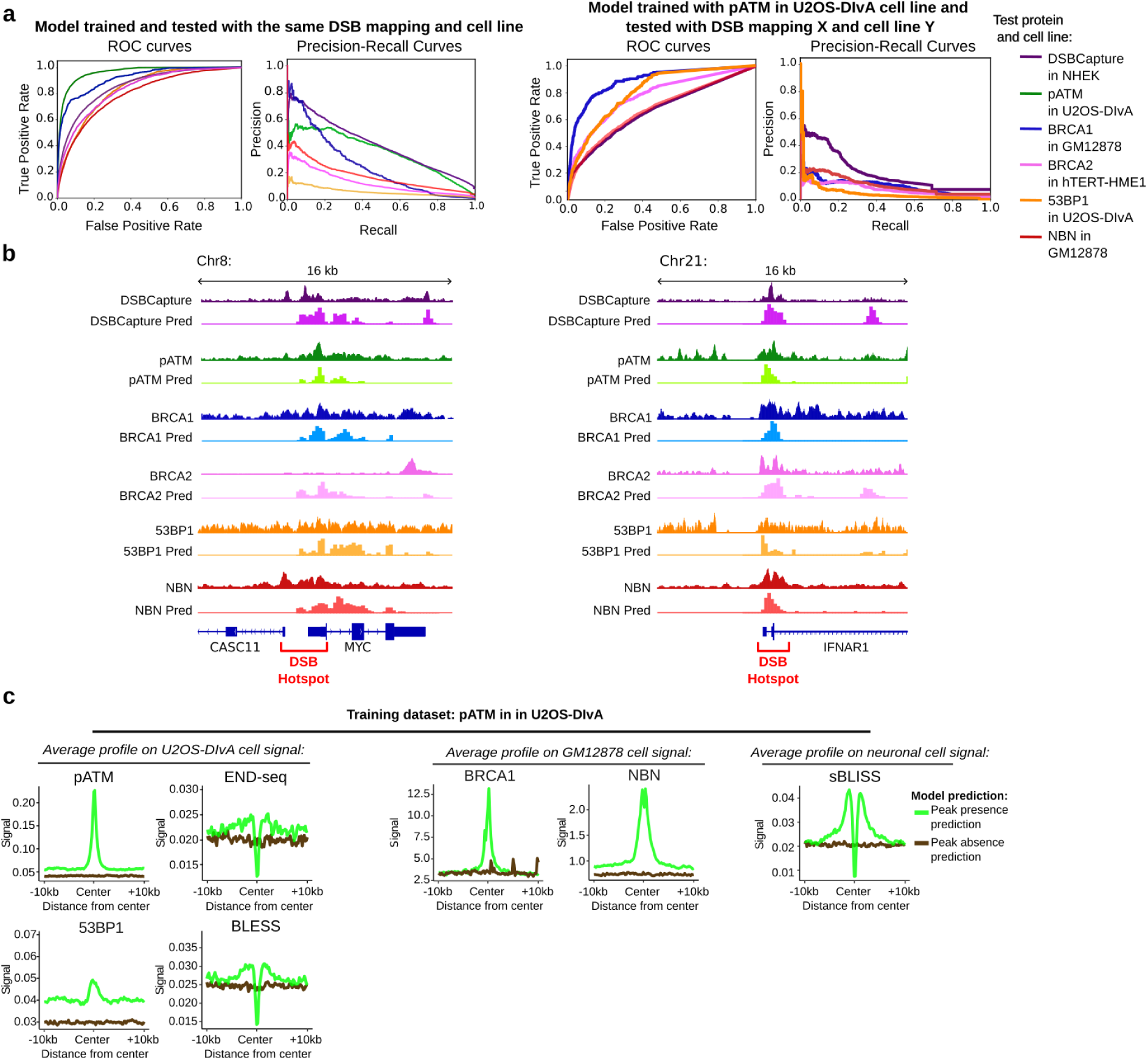
DNA sequence is a determinant of endogenous DSB frequency. **a)** Left: Receiver operating characteristic (ROC) curves and corresponding precision-recall (PR) curves of deep learning models trained and tested on the same DSB mapping method (either pATM, BRCA1, BRCA2, 53BP1, or NBN ChIP-seq, or DSBCapture) and the same cell line (either U2OS, NHEK, GM12878, or hTERT-HME1). U2OS-DIvA cells are derived from bone cancer, NHEK cells are epidermal keratinocytes, GM12878 cells are lymphoblastoid cells, and hTERT-HME1 cells are mammary epithelial cells. pATM stands for ‘phosphorylated ATM’. We trained the DeepSea network for binary classification (peak versus no peak). Right: ROC and PR curves of models trained on U2OS-DIvA pATM data and tested on other experiments and cell lines. **b)** Genome browser of two different regions showing pATM, DSBCapture, BRCA1, BRCA2, 53BP1, and NBN ChIP-seq data and their corresponding deep learning predictions. On the left side, the region around the *MYC* gene (MYC proto-oncogene; chromosome 8) is shown. On the right side, the region around *IFNAR1* gene (Interferon Alpha And Beta Receptor Subunit 1; chromosome 21) is displayed. **c)** Average profiles of U2OS-DIvA cell line DSB experiments (DSBCapture and ChIP-seq) depending on predicted peak presence vs absence. Here, a model was trained using pATM peaks from U2OS-DIvA cells. Average profiles of other cell lines and DSB experiments depending on predicted peak presence vs absence, using the model trained with pATM peaks from U2OS-DIvA cells.

**Figure S1:**
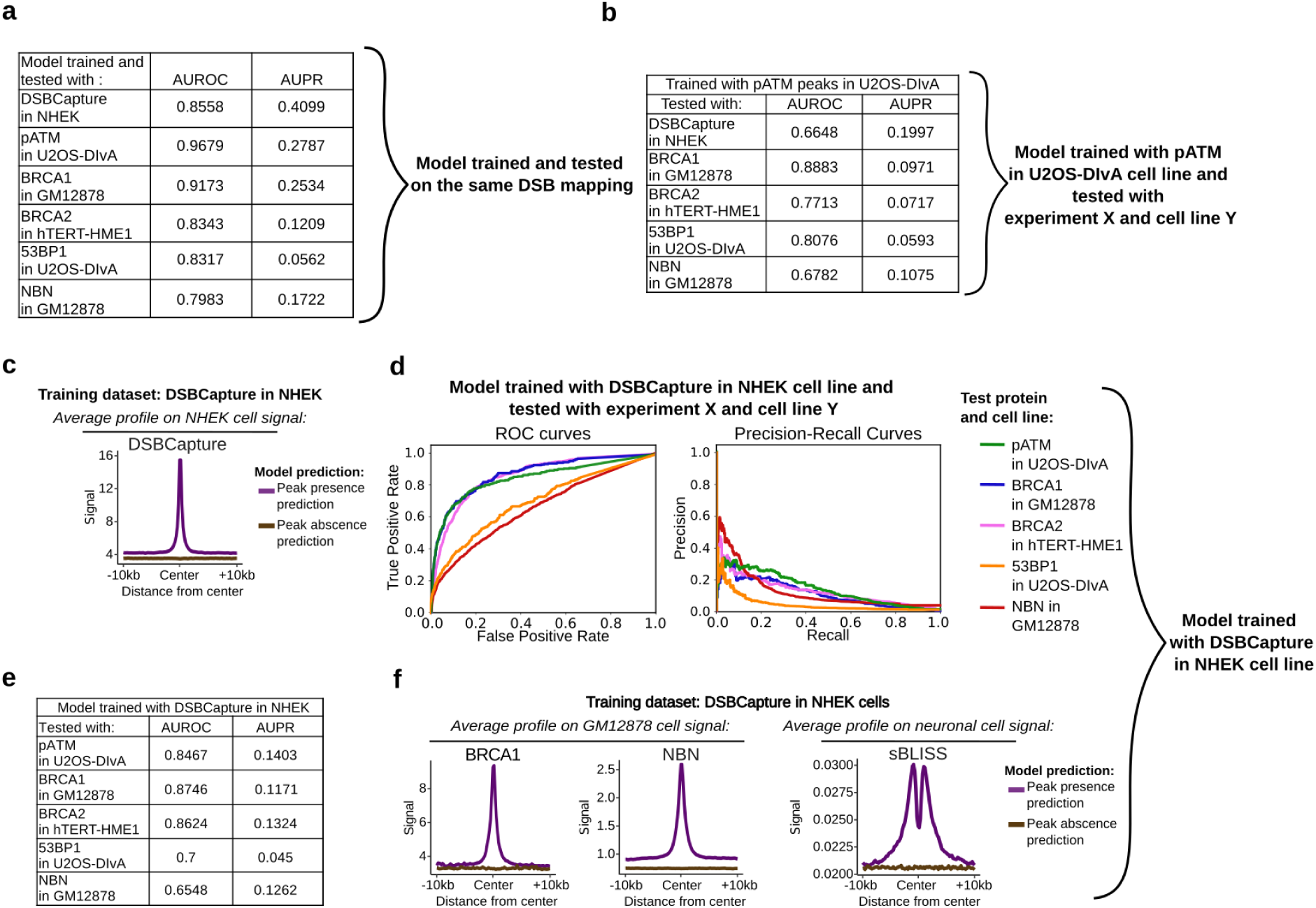
Prediction performance of models trained from DSB mapping data. **a)** Summary table of area under the ROC (AUROC) and area under the PR (AUPR) curves from Fig. 2a, left. The curves were computed from a test set of sequences (*n=120,000*), which was not used for model training. **b)** Summary table of corresponding AUROC and AUPR curves from Fig. 2a, right. **c)** Average profiles of NHEK DSBCapture experiment depending on predicted peak presence vs absence. Here, a model was trained using DSBCapture peaks from NHEK cells. The DeepSea network was used for binary classification (peak versus no peak). **d)** Receiver operating characteristic (ROC) curves and corresponding precision-recall (PR) curves of NHEK DSBCapture-trained model predictions, which were assessed on other experiments (53BP1, BRCA1, BRCA2, NBN, and pATM ChIP-seq) and cell lines (DIvA, GM12878, and hTERT-HME1). DIvA cells are from bone cancer, GM12878 cells are lymphoblastoid cells, and hTERT-HME1 cells are mammary epithelial cells. pATM stands for ‘phosphorylated ATM’. **e)** Summary table of corresponding area under the ROC (AUROC) and area under the PR (AUPR) curves from Fig. S2d. **f)** Average profiles of other cell lines and DSB experiments depending on predicted peak presence vs absence, using a model trained on DSBCapture peaks from NHEK cells.

To further assess the role of DNA as a determinant of endogenous DSB hotspots independently of the cell type, we used a model trained with pATM ChIP-seq data in U2OS-DIvA, to predict various DSB mapping experiments in other cell lines. The model showed great performances for BRCA1 in GM12878 cells (AUROC=0.89, AUPR=0.10; Fig. 2a, right side, and S1b). Similar results were found for BRCA2 in hTERT-HME1 cells^34^ and NBN in GM12878 cells, as well as for DSBCapture in NHEK cells^35^. Furthermore, using average profiles (Fig. 2c), we found high enrichment of BRCA1 and NBN in GM12878^36^, as well as high enrichment of sBLISS in neurons at predicted DSB peaks^37^. In addition, when using a model trained with the DSBCapture dataset in NHEK cells (keratinocytes)^35^, we also observed good predictions for several DSB mapping experiments in other cell lines (Fig. S1c-f).

Deep learning revealed that endogenous DSB frequency is predictable from, and thus strongly influenced by the DNA sequence. Therefore, variants, such as SNPs, that disrupt the DNA sequence could affect the frequency of endogenous DSBs and their repair. We hypothesized that such DSB SNPs (dsbSNPs) may be widespread along the genome and may play a role in complex genetic diseases.

### Mapping of dsbSNPs by allelic imbalance

Using allelic imbalance, we mapped DNA double-strand break SNPs (dsbSNPs) using several ChIP-seq experiments of DSB repair proteins performed under untreated conditions in U2OS-DIvA cells^29,30,38^. In addition to pATM ChIP-seq (Collins et al, in prep.), we analysed ChIP-seq data of four additional repair proteins (53BP1, SETX, RPA, and Lig4). The analyzed SNPs correspond to sites where endogenous DSBs frequently occur.

In total, the U2OS-DIvA cell genome contained 469,429 heterozygous SNPs. Among these, 1.3% were identified as dsbSNPs (6,013 unique dsbSNPs: 5,925 from pATM, 721 from 53BP1, 598 from SETX, 243 from RPA, 43 from Lig4 ChIP-seq datasets). Genomic annotations revealed a considerable enrichment of dsbSNPs in promoter regions (<1kb from TSS) compared to control SNPs (“ctrlSNPs”) randomly drawn from the 1000 genomes project with a MAF>10% (40.3% vs. 2.1%, FC=19, p<1×10^−16^) and in the 1st introns (0.38% vs. 0.07%, FC=5.7, p=2.9×10^−9^) (Fig. 3a). Focusing on pATM, the most general marker of DSBs, we compared the experimentally measured pATM effect sizes (ESs) with previous deep learning predictions, which revealed a higher predicted absolute ES for pATM dsbSNPs with higher experimental ES, confirming the predictions and validating our dsbSNP mapping (Fig. 3b). To assess the reproducibility of dsbSNP mapping, we investigated whether the same dsbSNPs could be identified from ChIP-seq experiments against different repair proteins. Although we did not dispose of as much data as for pATM, we found that a significant fraction of dsbSNPs were reproducible between 2 proteins (13.9%) or 3 proteins (3.7%) (Fig. S2a), reinforcing the reliability of dsbSNP mapping.

**Figure 3:**
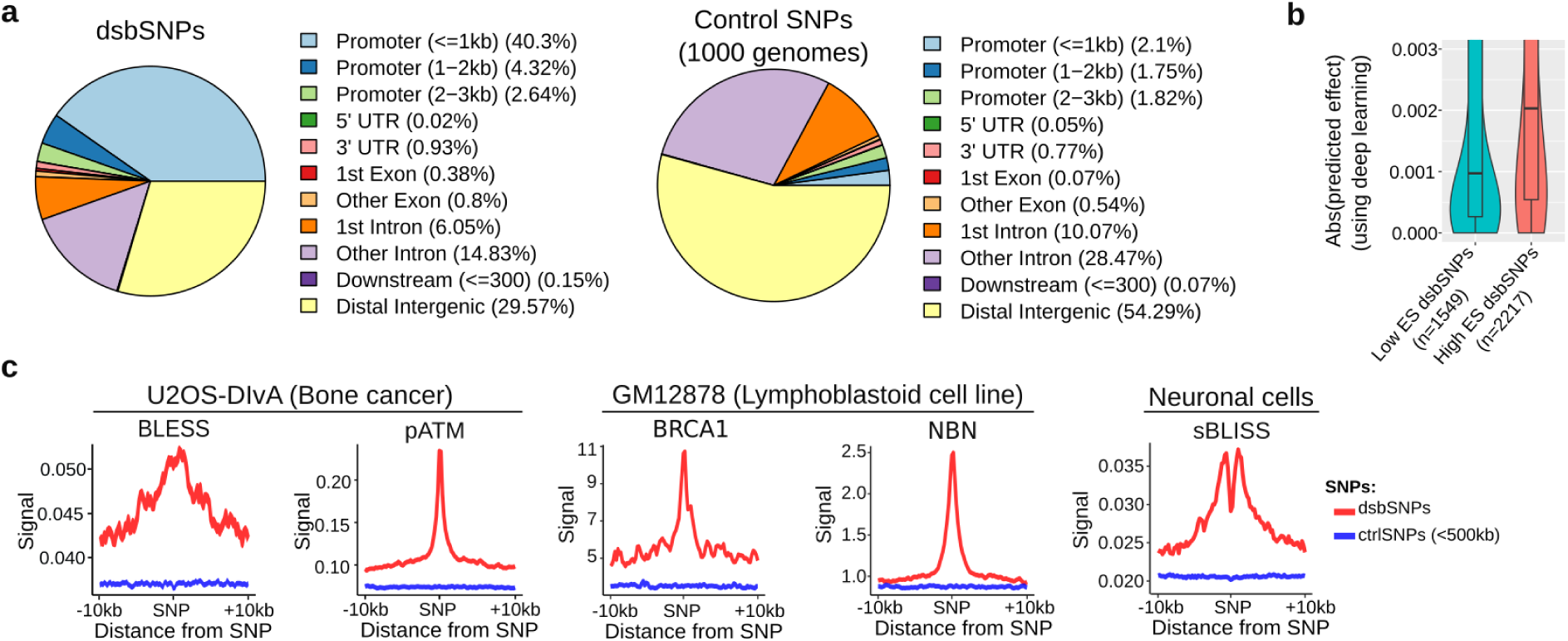
Genome-wide mapping of SNPs affecting DSB frequency (dsbSNPs) in DIvA cells. **a)** Genic annotations of dsbSNPs (*n=*6013) and of control SNPs (*n=*6013). Control SNPs were randomly drawn from the 1000 Genomes Project with a minor allele frequency (MAF)>10%. **b)** Absolute value of deep learning predicted pATM SNP effect for pATM dsbSNPs with high effect size (abs(effect size) > 1), as compared to pATM dsbSNPs with low effect size (abs(effect size) < 0.5). The p-value was computed using a two-sided nonparametric Wilcoxon rank-sum test. **c)** Average profiles of DSB mapping (BLESS, sBLISS, and pATM, BRCA1, and NBN ChIP-seq) at dsbSNPs in cancer (U2OS-DIvA), immune cells (GM12878), and neuronal cells. Here, control SNPs “ctrl SNPs (<500 kb)” were randomly drawn from the 1000 Genomes Project with a MAF>10% and from less than 500 kb from dsbSNPs.

**Figure S2:**
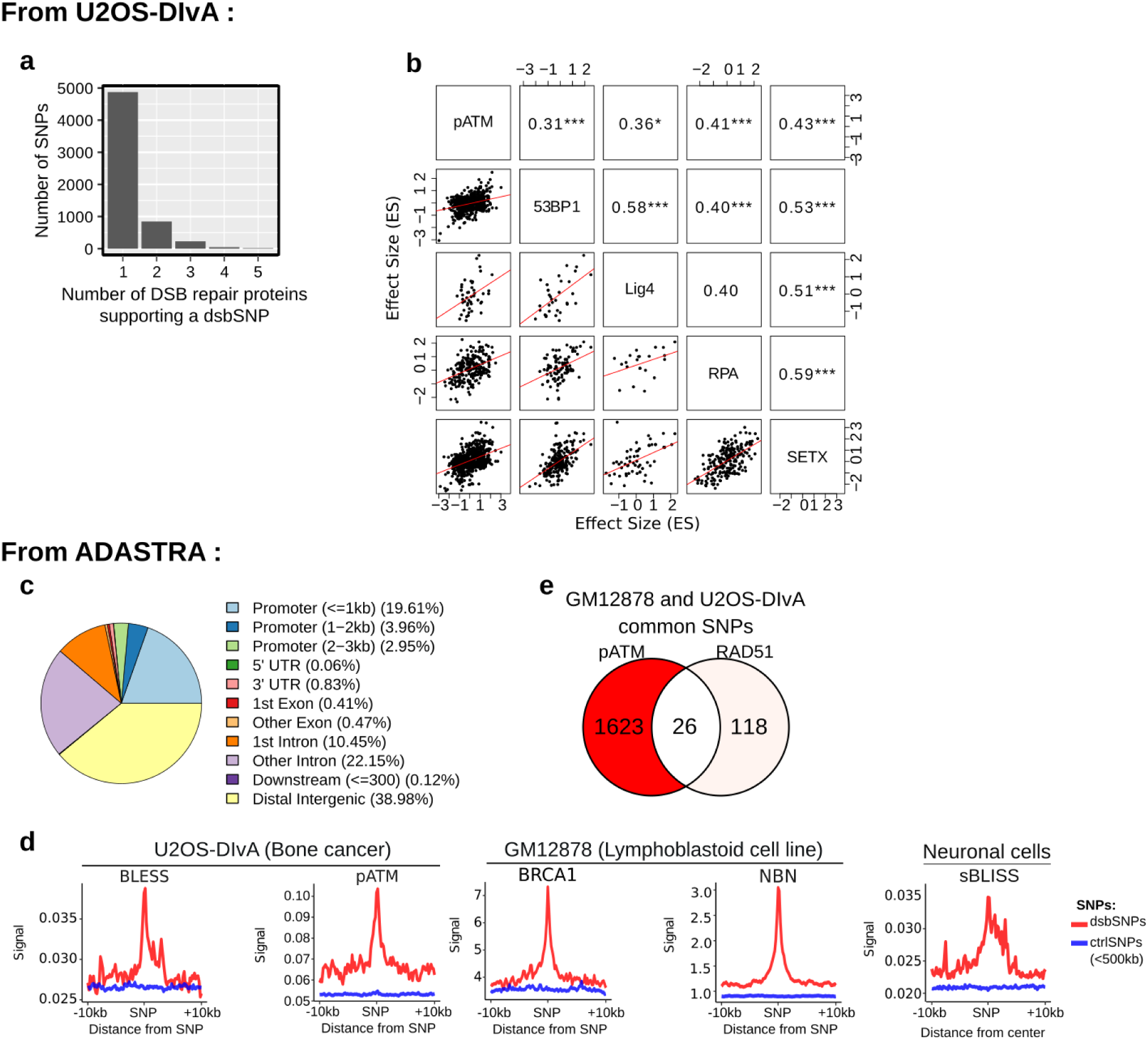
SNPs affecting DSB frequency (dsbSNPs) in U2OS-DIvA cells and from the ADASTRA database restricted to SNPs affecting DNA repair protein binding. **a)** Number of DSB repair proteins supporting a dsbSNP in U2OS-DIvA cells which corresponds to the number of DSB repair proteins whose binding is in allelic imbalance for a given dsbSNP. **b)** Scatter plots and Pearson correlations between observed SNP effect sizes from different DSB repair proteins in U2OS-DIvA cells. For a pair of proteins, only the dsbSNPs that are in allelic imbalance in both proteins are shown. * p<0.05, ** p<0.01, *** p<0.001. **c)** Genic annotation of ADASTRA dsbSNPs. **d)** Average profiles of DSB mapping (BLESS, sBLISS, and pATM, BRCA1, and NBN ChIP-seq) at ADASTRA dsbSNPs (*n=1598*), in cancer (U2OS-DIvA), neurons, and immune cells (GM12878). dsbSNPs were compared to control SNPs (*n=1598*). Here, control SNPs “ctrl SNPs (<500 kb)” were randomly drawn from the 1000 genomes project with a MAF>10% and from less than 500 kb from dsbSNPs. **e)** Venn diagram showing the overlap between pATM SNPs mapped from our U2OS-DIvA cells and RAD51 SNPs mapped from GM12878 cells from ADASTRA. Here, we only kept SNPs that were genotyped as heterozygous SNPs in both U2OS-DIvA and GM12878 cells.

**Figure S3:**
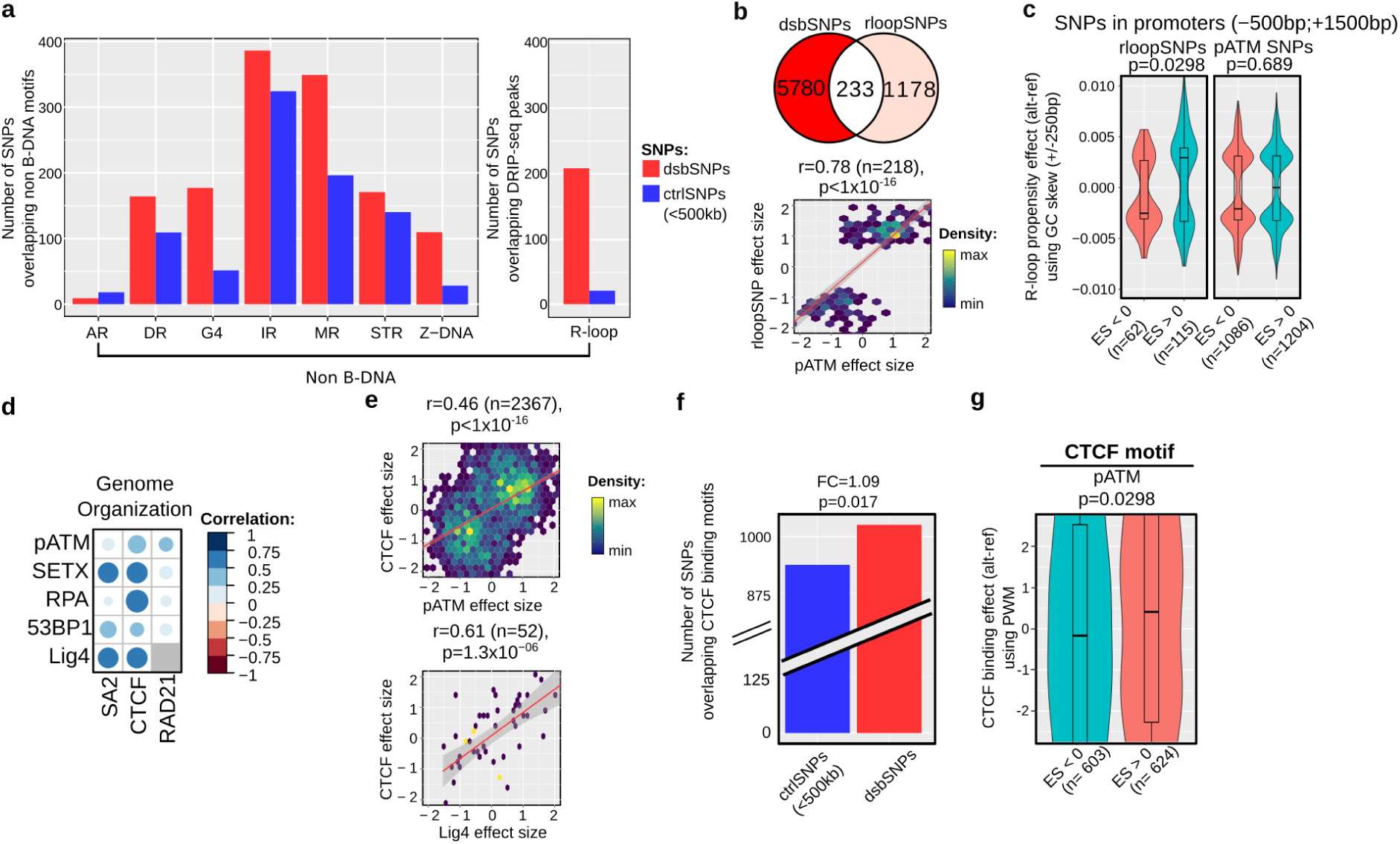
Non-B DNA and 3D genome determinants of DSBs. **a)** Enrichment of dsbSNPs (as compared to control SNPs) at non-B DNA structures: A-phase repeats (AR), directed repeats (DR), G-quadruplexes (G4s), inverted repeats (IRs), mirror repeats (MRs), R-loops (three-stranded RNA:DNA hybrids), short tandem repeats (STRs), and Z-DNA. Here, control SNPs “ctrl SNPs (<500 kb)” were randomly drawn from the 1000 genomes project with a MAF>10% and from less than 500 kb from dsbSNPs. **b)** Venn diagram of dsbSNPs and rloopSNPs, and scatter plot of dsbSNP effect size and rloopSNP effect size. **c)** Effect of rloopSNPs and pATM dsbSNPs on R-loop propensity (measured by GC skew), for SNPs with effect size > 0 as compared to SNPs effect size < 0. The R-loop propensity effect was defined as the difference in GC skew between the alternative and reference alleles. The p-values were computed using two-sided nonparametric Wilcoxon rank-sum tests. **d)** Correlation heatmap between SNP effect sizes (ESs) from different DSB repair proteins and SNP effect sizes from 3D genome proteins (CTCF, SA2, and RAD21). **e)** Scatter plot of effect sizes between pATM and CTCF SNP ESs, and between Lig4 and CTCF SNP ESs. **f)** CTCF binding motif enrichment at dsbSNPs vs control SNPs. **g)** Effect of dsbSNPs on CTCF binding for dsbSNPs with effect size > 0 as compared to dsbSNPs effect size < 0. CTCF binding effect was computed as the difference of the CTCF motif position weighted matrix (PWM) score between the alternative and reference alleles. The p-value was computed using a two-sided nonparametric Wilcoxon rank-sum test.

To investigate the association between dsbSNPs from different repair proteins, we measured the pairwise correlation between the protein binding allelic effect sizes (Fig. S2b). As expected, repair proteins SNP ES were positively correlated. Among these correlations, SETX (R-loop helicase) and RPA (single-strand DNA-binding factor) displayed a high positive correlation (r=0.59), in agreement with the reported functional link between R-loop and resection^39,30^. Similarly, 53BP1 and Lig4, two proteins involved in NHEJ repair^38^, also showed a high positive correlation (r=0.58). In addition, by calculating average profiles, we confirmed that dsbSNPs precisely colocalized with endogenous DSBs as mapped by BLESS and pATM ChIP-seq in U2OS DIvA cells (Fig. 2c, left) (here, control SNPs were the same as above, but drawn from less than 500 kb from dsbSNPs). Furthermore, the same results were found with BRCA1 and NBN ChIP-seq in GM12878 cells^36^ (Fig. 2c, middle) or with sBLISS in neuronal cells^37^ (Fig. 2c, right).

In addition, we replicated our analyses to identify dsbSNPs using the ADASTRA database of allele-specific variants, which comprises a large set of SNPs identified from publicly available ChIP-seq experiments in various cell lines. In total, using 3,677,218 heterozygous SNPs from the database, we identified 1,676 different dsbSNPs (0.04%): 1,138 from RAD51 binding, 116 from NBN, 421 from BRCA2, and 14 from BRCA1 binding (Fig. S2c). Similarly to the dsbSNPs mapped from data acquired in U2OS-DIvA cells, these dsbSNPs were highly enriched in the promoter regions (<1kb from TSS) (19.61% vs. 2.1%, FC=9.3 p<1×10^−16^) and in the first introns (0.41% vs. 0.07%, FC=6.2, p=1.6×10^−10^) as compared to control SNPs (Fig. S2c). By calculating average profiles, we confirmed that dsbSNPs identified using the ADASTRA database colocalized with endogenous DSBs (Fig. S2d). When we overlapped pATM SNPs from U2OS-DIvA cells with RAD51 SNPs identified in GM12878 cells available in the ADASTRA database, we found that 18% of RAD51 SNPs (26 out of 144) were also found as pATM SNPs in U2OS-DIvA cells, further validating our U2OS-DIvA dsbSNPs (Fig. S2e).

Together, these data uncovered a novel class of SNPs, which we termed dsbSNPs, associated with DSB frequency, representing at least 1.3% of SNPs in the genome, and which are mostly found in non-coding elements, such as promoters and introns. Importantly, we are able to identify dsbSNPs with a very high resolution of ∼250 bp, as compared to GWAS SNPs which were identified at ∼10-20 kb resolution in European populations^11^. To create a large dsbSNP dataset for further downstream analyses, we then decided to merge the dsbSNPs mapped from our data (in U2OS-DIvA cells) with the dsbSNPs mapped from the ADASTRA database, yielding a joint set of 7,643 unique dsbSNPs.

### Non-B-DNA and 3D genome determinants

Alternative DNA conformations, deviating from the usual B-form double helix, can naturally form in the genome and have previously been hypothesized to be associated with genome instability and DSBs. These non-B DNA structures include: (i) G-quadruplexes (G4s) where DNA folds into four-stranded structures, often at G-rich regions; (ii) Z-DNA which corresponds to a left-handed DNA helix, compared to the usual right-handed B-DNA; (iii) R-loops also denoted RNA:DNA hybrids, which are three-stranded structures that form when RNA hybridizes to a DNA strand, displacing the other DNA strand; (iv) Repeat structures encompassing A-phase repeats (AR), directed repeats (DR), inverted repeats (IRs), mirror repeats (MRs) and short tandem repeats (STRs). We were able to look for enrichment of dsbSNPs in predicted non-B DNA structures^41^. We found that dsbSNPs were especially enriched in predicted G4 (FC=3.4, p=2×10^−17^) and Z-DNA (FC=3.9, p=1×10^−12^) forming sequences (Fig. S3a). Moreover, we mapped R-loops in U2OS-DIvA cells by DRIP-seq^29,42^ and found a high enrichment of dsbSNPs at R-loop peaks (FC=9.9, p=2.17×10^−39^) (Fig. S3a).

R-loops have been repeatedly linked to genome instability, and multiple studies have suggested that these transcriptionally-induced non-B DNA structures could trigger DSBs, for instance by impeding the progression of the replication fork. However, R-loops (or RNA:DNA hybrids) also arise in response to DSB production^27,28,41^, making it difficult to establish whether R-loops are a cause or consequence of DSB. To further explore this link, we identified R-loop SNPs (rloopSNPs) from the DRIP-seq dataset using allelic imbalance. In total, 233 out of 1,178 (16.5%) rloopSNPs were dsbSNPs (Fig. S3b). We observed a strong positive correlation between pATM SNP and rloopSNP effect sizes (ESs) with a Pearson correlation of 0.78 (p<1×10^−16^; Fig. S3b), highlighting, as expected, a tight link between R-loop formation and DSB frequency^39^. Furthermore, since R-loops are strongly regulated by the DNA sequence and are more susceptible to form and to be stabilized on DNA G stretches^44^, we used rloopSNPs and dsbSNPs to examine causality by looking at GC skew, an approach similar to the one used in^45^. As expected, rloopSNPs at promoters are influenced by GC skew alteration (p=0.0298). In contrast, pATM SNPs were not (p=0.689) (Fig. S3c), strongly suggesting that the previously established correlation between R-loops and DSB may arise from the accumulation of R-loops at DSBs, rather than R-loops generating DNA damage, at least in promoter regions.

DSB generation in the genome has also been previously associated with chromosome architecture, particularly increased fragility of topologically associated domains (TADs) boundaries bound by CTCF^46,47^. To investigate the link with chromosome architecture, we have also mapped the allele-specific binding sites at SNPs in U2OS-DIvA cells for CTCF, cohesin subunits STAG2 (SA2), and RAD21. We found that most repair proteins showed ESs positively correlated with CTCF ESs, such as pATM (r=0.47, p<1×10^−16^; Fig. S3d and S3e) and Lig4 (r=0.61, p<1.3×10^−5^; Fig. S3d and S3e). As previously done to examine the causality between R-loop and DSB, we further investigated a potential causal effect of CTCF motifs on DSBs. For pATM SNPs, we first found a slight enrichment at CTCF binding motifs (FC=1.09, p=0.017; Fig. S3f). Most notably, for pATM SNP alleles with a positive ES, we observed a slightly higher effect on CTCF motifs (p=0.0298, Fig. S3g), strengthening the previously proposed causal effect of CTCF binding on DSB frequency.

Altogether, our data suggest that the established correlation between R-loop and DSB may not originate from the ability of R-loop to induce DSB, but rather from R-loops accumulating after DSB formation. Additionally, our data further reinforce the previously proposed susceptibility of CTCF-bound as being prone to DSB. Accordingly, we show that SNPs disrupting CTCF motifs may affect genome instability.

### dsbSNPs alter regulatory element activity and gene expression in normal tissues and cancers

The annotation of dsbSNPs indicated a potential association with regulatory elements, especially promoters (Fig. 3a and Fig. S2c). In agreement, we found almost 2.58-fold enrichment of H3K4me3, 1.55-fold enrichment of H3K27ac and 2.02-fold enrichment of ATAC-seq (measuring chromatin accessibility and performed in U2OS-DIvA cells, Collins et al, in prep.) at dsbSNPs compared to control SNPs (<500 kb) (Fig. 4a). The effect sizes between pATM SNPs and H3K4me3 (R=0.39, p<1×10^−16^) or H3K27ac (R=0.44, p<1×10^−16^) SNPs were positively correlated (Fig. 4b), confirming the strong association with regulatory elements. Similarly, we found a positive correlation between pATM SNP ES and ATAC SNP ES (R=0.76, p<1×10^−16^) (Fig. 4b).

**Figure 4:**
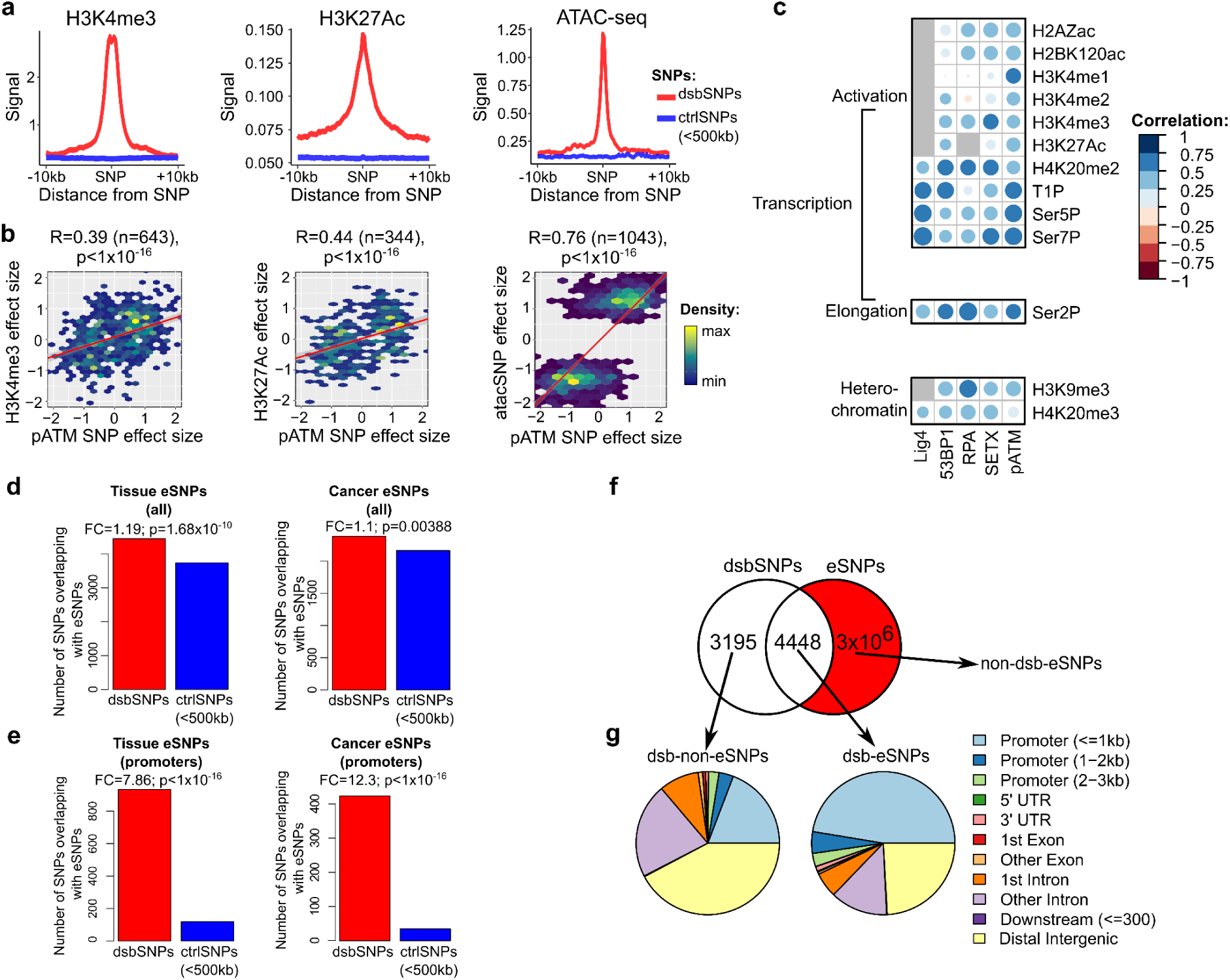
Association of dsbSNPs with regulatory elements and gene expression. **a)** Average profile of H3K4me3, H3K27ac ChIP-seq and ATAC-seq at dsbSNPs compared to control SNPs. Here, control SNPs “ctrl SNPs (<500 kb)” were randomly drawn from the 1000 genomes project with a MAF>10% and from less than 500 kb from dsbSNPs. **b)** Scatter plot of pATM SNP effect size and corresponding H3K4me3, H3K27ac, and ATAC-seq SNP effect sizes. **c)** Correlation heatmap between SNP effect sizes from different DSB repair proteins and SNP effect sizes from different chromatin marks. **d)** Number of dsbSNPs (as compared to control SNPs) overlapping normal tissue expression SNPs (eSNPs), and overlapping germline cancer eSNPs. **e)** Number of dsbSNPs at promoters (as compared to control SNPs) overlapping normal tissue eSNPs, and overlapping germline cancer eSNPs. **f)** Venn diagram of overlap between dsbSNPs and eSNPs. **g)** Genic annotations of dsbSNPs overlapping eSNPs (“dsb-eSNPs”) and of dsbSNPs not overlapping eSNPs (“dsb-non-eSNPs”).

**Figure S4:**
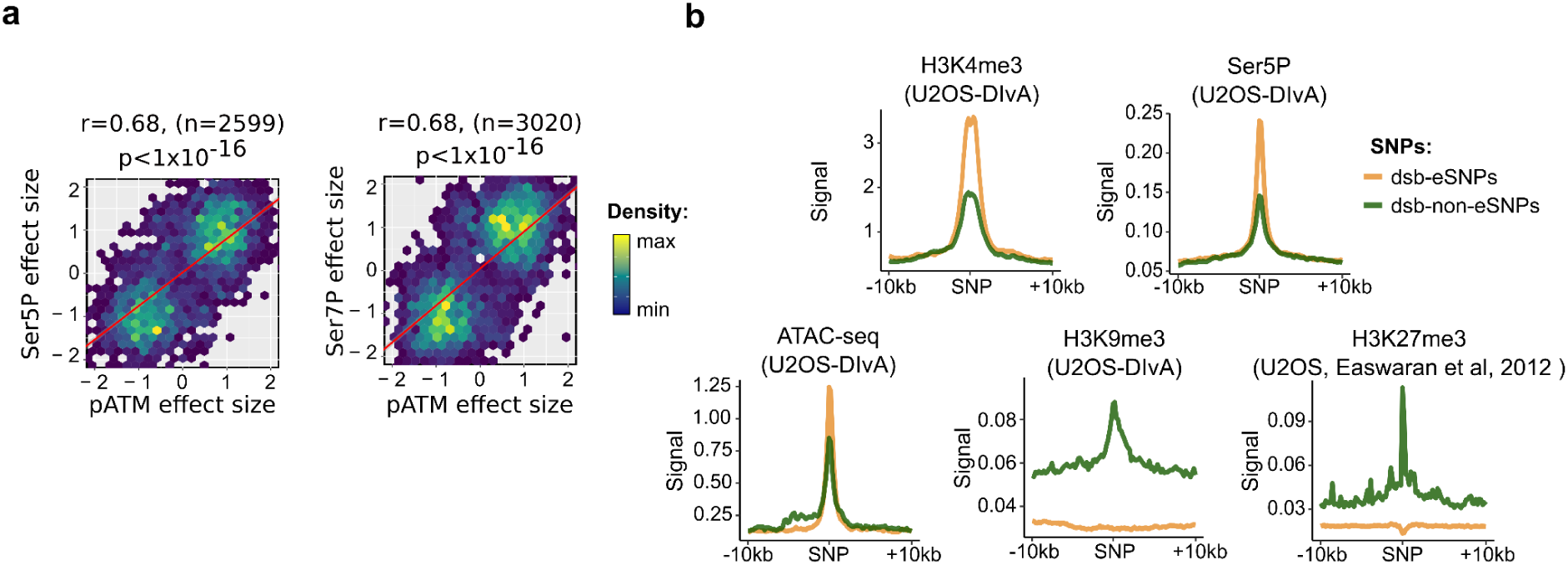
Link between dsbSNPs and chromatin marks. **a)** Scatter plot of effect sizes between repair protein pATM and transcription initiation marks RNAPII Ser5p and Ser7p. **b)** Average profiles of histone mark ChIP-seq^48^, and ATAC-seq at dsb-eSNPs and dsb-non-eSNPs.

Transcription has been hypothesized as either causing or requiring DSB; therefore, we sought to study the link between transcription and DSB. Our datasets previously produced in U2OS-DIvA cells also allowed us to map by allelic imbalance SNPs both for histone marks and RNA polymerase binding, which are proxies for transcriptional activity. First, we looked at transcription initiation by examining RNA polymerases modified on Serine 5, Serine 7, or Tyrosine 1 (RNAPII Ser5P, RNAPII Ser7P, RNAPII T1P) that are markers of transcription initiation^49^. We observed that pATM SNP effect sizes (ESs) were positively correlated with the SNP ESs of RNAPII Ser5P, RNAPII Ser7P, and RNAPII T1P (r=0.68, p<1×10^−16^; r=0.68, p<1×10^−16^; r=0.63, p=3.6×10^−18^, resp.; Fig. 4c and S4a). Such a positive relation between DNA repair proteins and RNAPII modifications could be explained either by transcriptional activity that could generate DSBs^50^, or by transcription initiation requiring DSBs^51,52^. Next, we focused on transcription elongation by looking at RNA polymerase modified on Serine 2 (RNAPII Ser2P) which corresponds to a modification of the C-terminal domain of RNAPII, and found that the SNP ESs were indeed correlated (r=0.71, p=4.6×10^−30^; Fig. 4c). This further strengthens the previously observed link between homologous recombination, resection and transcriptional activity.

Given the strong association between dsbSNPs and regulatory elements, we then investigated the association between dsbSNPs and SNPs associated with gene expression (expression SNPs, eSNPs) previously mapped in physiological tissues by the GTEx consortium^14^. We detected a 1.19-fold enrichment at normal tissue eSNPs compared to control SNPs (p=1×10^−10^) (Fig. 4d). Similarly, we mapped dsbSNPs to eSNPs identified in cancer (cancer eSNPs) from the ICGC consortium and found a 1.1-fold enrichment compared to control SNPs (p=0.004, Fig. 4d). More interestingly, when focusing only on eSNPs located at promoters, we detected a 7.9-fold enrichment of dsbSNPs over normal tissue eSNPs (vs. control SNPs; p<1×10^−16^, Fig. 4e) and a 12.3-fold enrichment of dsbSNPs over cancer eSNPs (vs. control SNPs; p<1×10^−16^; Fig. 4e), revealing a tight link between dsbSNPs and transcription, especially in cancer.

In light of the strong association between dsbSNPs and transcription, we next decided to distinguish between dsbSNPs that overlapped eSNPs (“dsb-eSNPs”) and dsbSNPs that did not overlap eSNPs (“dsb-non-eSNPs”) to better characterize the link between DSB and transcription. Out of 7,643 dsbSNPs, there were 4,448 (58%) dsb-eSNPs and 3,195 (42%) dsb-non-eSNPs (Fig. 4f). dsb-eSNPs were mainly found in promoters (47.39%), whereas dsb-non-eSNPs were mainly found in distal intergenic regions (42.25%) (Fig. 4g). In agreement with the ability of dsb-eSNPs to regulate gene expression, dsb-eSNPs were enriched in RNAPII Ser5P and H3K4me3, and in ATAC-seq signals (Fig. S4b). Conversely, dsb-non-eSNPs showed an enrichment in both H3K27me3 and H3K9me3 signals that are heterochromatin marks and lower levels of promoter active mark H3K4me3 and ATAC-seq signals (Fig. S4b).

It is worth mentioning that contrary to dsbSNPs that are mapped at a very high resolution (∼250bp), eSNPs are usually mapped at a very low resolution (∼10-20 kb in Europeans) and therefore these results are to be interpreted with caution. However, altogether these results further support the association between dsbSNPs and active chromatin SNPs as well as non-coding SNPs that regulate gene expression (eSNPs). Conversely, SNPs acting independently of transcription (dsb-non-eSNPs, 42% of SNPs) were associated with heterochromatin marks and lower levels of promoter active mark.

### Transcription factor determinants of the selection of the repair pathway

Since certain TF motifs are strong predictors of DSB frequency^25^, we next evaluated the enrichment of dsbSNPs at TF motifs and detected several enriched motifs, such as TFEB (FC=16.5, p=1.5×10^−15^) and TCFL5 (FC=9.7, p<1×10^−16^) (Fig. 5a). As previously, we used TF motifs to decipher whether transcription is a cause or consequence of DSB. For pATM SNPs, we did not observe any effect on TF motif score alteration (p=0.747; Fig. 5b), suggesting that transcription activation is not causal to DSB production. Yet, strikingly, we found a strong positive relation between TF binding and RAD51 SNP ES (p=1×10^−16^; Fig. 5b), revealing that SNPs increasing TF binding can promote the homologous recombination (HR) DSB repair pathway, as RAD51 is a key marker of HR. To confirm this result, we investigated the link with 53BP1, which is known to favor the NHEJ repair pathway. Accordingly, we observed a negative relation between TF binding and 53BP1 SNP ES (p=7×10^−16^; Fig. 5b), revealing that SNPs increasing TF binding, while promoting HR usage, also inhibit NHEJ repair. When looking specifically at the most enriched TF motifs (BHLHE40 and TCFL5; Fig. 5a), we found similar results but with much stronger effects (Fig. 5c and 5d). Moreover, in agreement with TF motif analysis, dsb-eSNPs, which are involved in transcription, showed high RAD51 level linked to HR, while dsb-non-eSNPs showed instead high 53BP1 and Lig4 signals associated with NHEJ (Fig. 5e).

**Figure 5:**
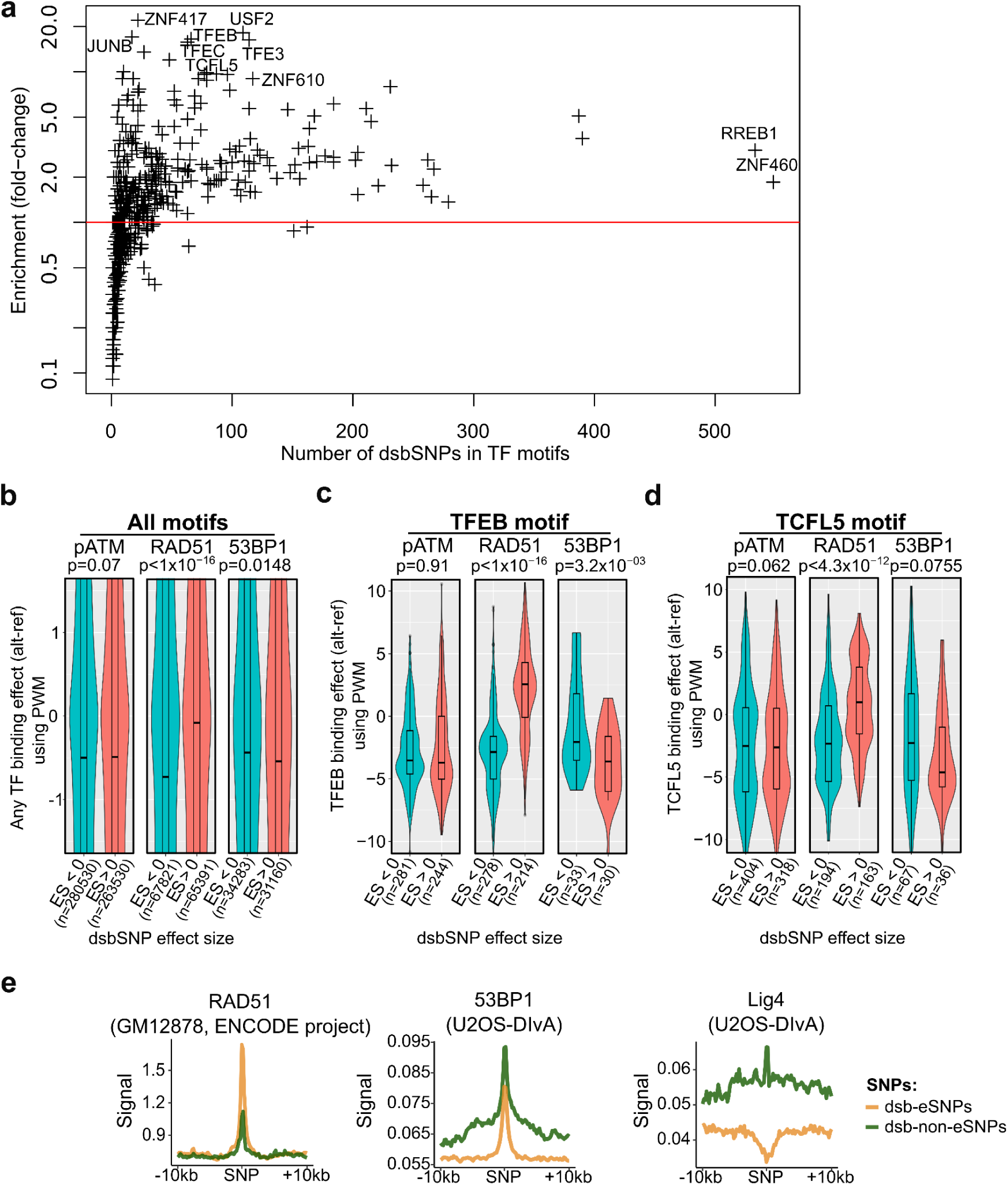
Transcription factor determinants of DSB repair pathway selection. **a)** Enrichment of dsbSNPs (vs control SNPs) at transcription factor (TF) DNA motifs. Motifs were mapped using position weight matrices (PWMs). Here, control SNPs were randomly drawn from the 1000 genomes project with a MAF>10% and from less than 500 kb from dsbSNPs. **b)** Effect of dsbSNPs (pATM, Rad51, and 53BP1) on all TF bindings for dsbSNPs with effect size > 0 as compared to dsbSNPs with effect size < 0. For a given TF, the binding effect was computed as the difference between the position weight matrix (PWM) scores for the alternative and reference alleles. The p-values were computed using two-sided nonparametric Wilcoxon rank-sum tests. **c)** Effect of dsbSNPs on BHLHE40 binding motifs for dsbSNPs with effect size > 0 as compared to dsbSNPs effect size < 0. The binding effects and p-values were computed as in d). **d)** Same as e) for TCFL5 binding motifs. **e)** Average profiles of DNA repair proteins at dsbSNPs overlapping eSNPs (“dsb-eSNPs”) and dsbSNPs not overlapping eSNPs (“dsb-non-eSNPs”).

**Figure S5:**
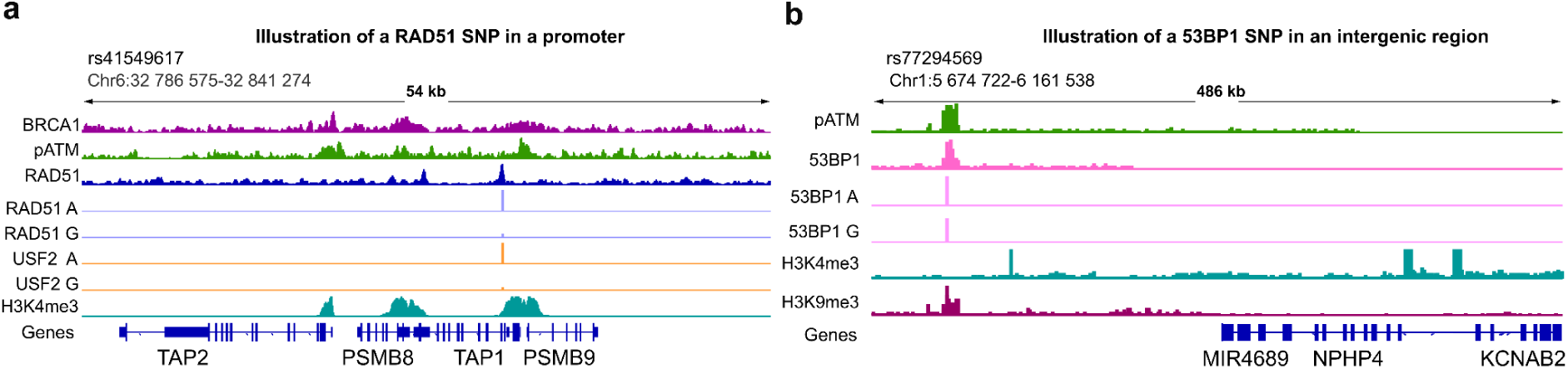
Genomic view of two dsbSNPs. **a)** Genome browser showing the SNP rs41549617, a RAD51 SNP located at the promoter of *TAP1* (*Transporter 1, ATP Binding Cassette Subfamily B Member*) gene. **b)** Genome browser displaying the SNP rs77294569, a 53BP1 SNP located in an intergenic region near *MIR4689* (MicroRNA 4689). Integrative Genome Viewer was used.

For illustration purposes, we then picked one RAD51 SNP (rs41549617) and one 53BP1 SNP (rs77294569) (Fig. S5a and S5b, resp.). Allele A of rs41549617 (as compared to allele G) was associated with higher RAD51 binding, as well as higher TF USF2 binding as measured by allelic imbalance, and was located at the active promoter mark H3K4me3. Conversely, allele A of rs77294569 (as compared to allele G) was associated with higher 53BP1 binding. The SNP showed no H3K4me3 signal, but instead a high signal of heterochromatin mark H3K9me3.

Altogether, our data suggest that while DSB formation correlates with transcription, transcriptional activity may not be causal to DSB formation. However, variants that disrupt transcription factor motifs affect DSB repair, notably the use of homologous recombination, suggesting that transcriptional activity contributes to determining the use of HR, as previously proposed^53^. Moreover, by regulating the selection of the repair pathway, such variants may influence the local mutation rate, since the HR pathway is intrinsically less error-prone than NHEJ^54^.

### dsbSNPs are associated with all classes of complex diseases

We then sought to assess whether dsbSNPs were associated with complex diseases. For this purpose, we analyzed the overlap of dsbSNPs with GWAS top associations from the EBI GWAS catalog (compiling 178,198 non-coding SNPs associated with at least one complex disease). For comparison purposes, we used two types of control SNPs from the 1000 genomes project with a MAF>10%, either randomly picked in the genome (“ctrlSNPs”), or randomly picked within a 200-500 kb distance from dsbSNPs (“ctrlSNPs (200-500kb)”) and eSNPs. The range from 200 kb to 500 kb was chosen to avoid any linkage disequilibrium while preserving a similar chromatin environment with the tested dsbSNPs. Among the 7,643 dsbSNPs, 386 mapped to known GWAS top associations (as compared to 175 ctrlSNPs and 210 ctrlSNPs (200-500kb)), corresponding to a strong enrichment (FC=1.84, p=2×10^−12^; FC=2.21, p=1.12×10^−18^; resp. Fig. 6a), suggesting a functional role of dsbSNPs in genetic diseases. Strikingly, dsbSNPs were twice as enriched at GWAS top associations than eSNPs (386/7,643 vs 187/7,643, FC=2.06, p=1.7×10^−16^, Fig. 6a).

**Figure 6:**
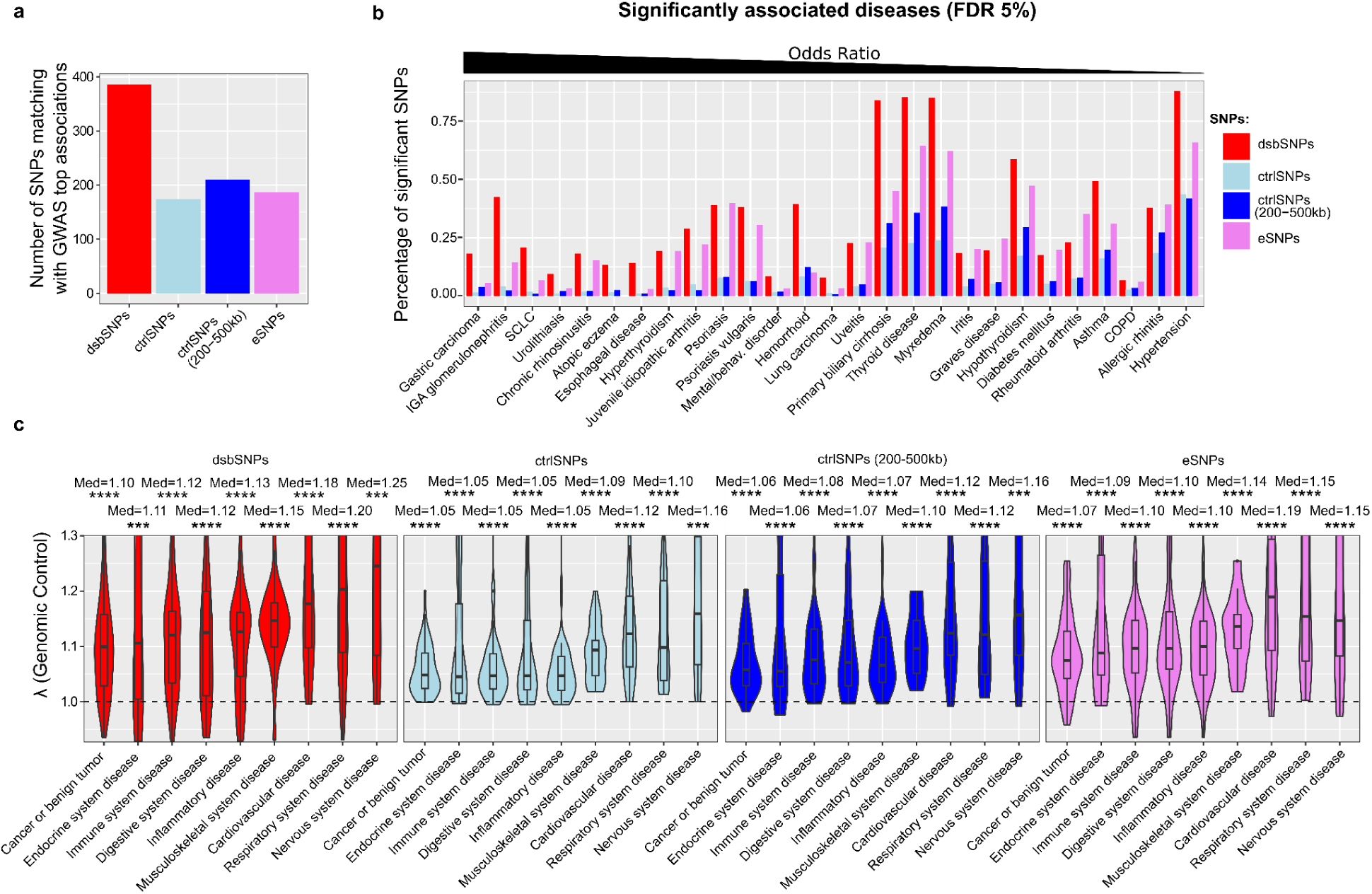
Association of dsbSNPs with complex genetic diseases. **a)** Overlap between dsbSNPs and GWAS top associations. The p-value was computed using Fisher’s exact test. Here, two types of control SNPs were randomly picked from the 1000 genomes project with a MAF>10%, either from the whole genome (“ctrlSNPs”), or within a 200-500 kb distance from dsbSNPs (“ctrlSNPs (200-500kb)”). **b)** Percentage of significant GWAS SNPs (p<10^−8^) for dsbSNPs as compared to control SNPs and eSNPs, for the significantly associated diseases (FDR=5%). Fisher’s exact test was used to test the significance of the association between dsbSNPs and significant GWAS SNPs. SCLC: squamous cell lung carcinoma; COPD: Chronic obstructive pulmonary disease. For some diseases, multiple GWAS studies were aggregated. **c)** For each disease class, genomic control factors λ for dsbSNPs, control SNPs, and eSNPs. Disease classes were defined by the GWAS Catalog^55^. The median (“Med”) is shown. P-values were computed using a Wilcoxon signed-rank test (H_0_: median=0). * p<0.05, ** p<0.01, *** p<0.001, **** p<0.0001.

**Figure S6:**
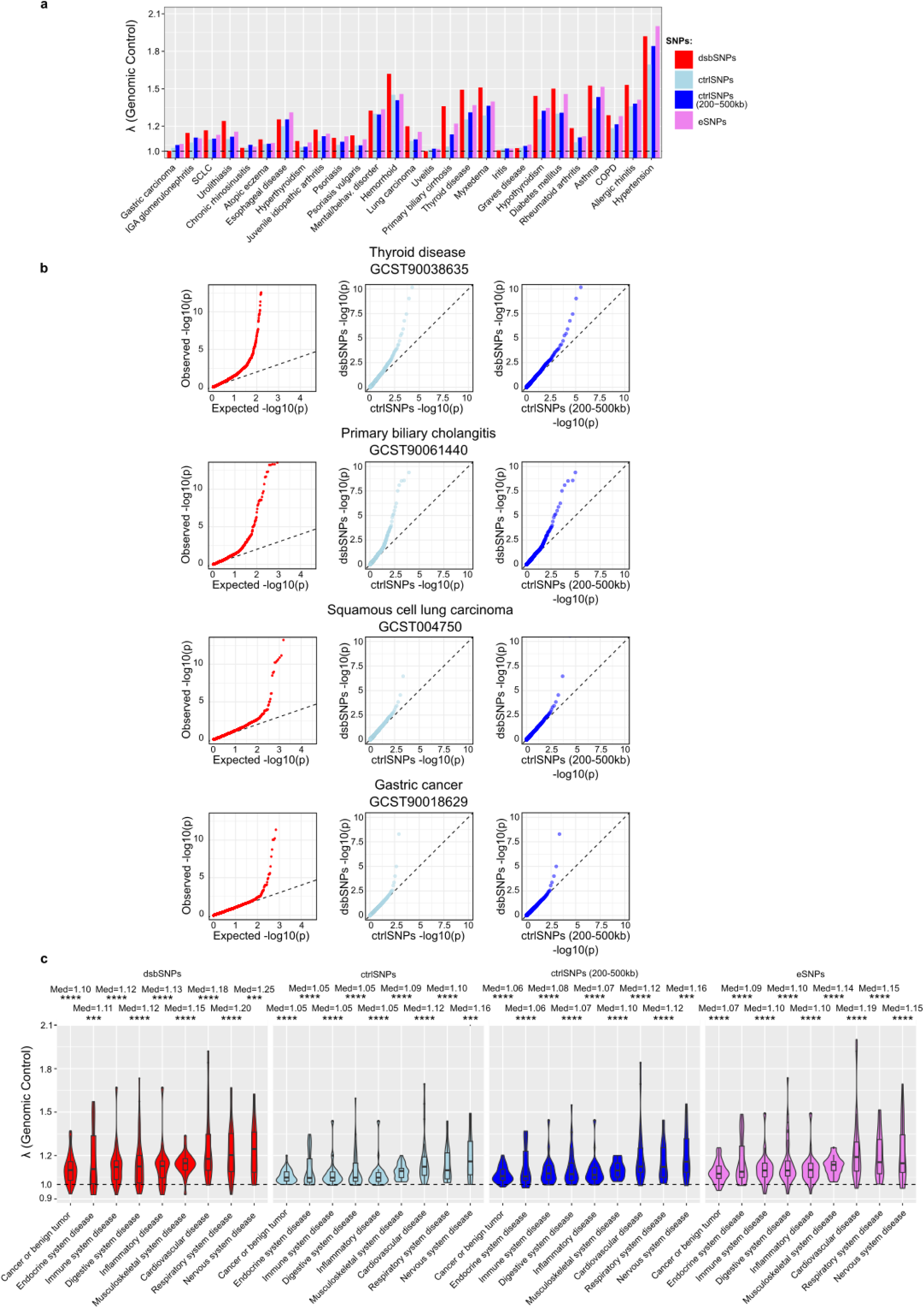
In-depth analysis of the genomic control. **a)** Genomic control λ for each disease tested from GWAS Catalog. Here, two types of control SNPs were randomly picked from the 1000 genomes project with a MAF>10%, either from the whole genome (“ctrlSNPs”), or within a 200-500 kb distance from dsbSNPs (“ctrlSNPs (200-500kb)”). SCLC: squamous cell lung carcinoma; COPD: Chronic obstructive pulmonary disease. For some diseases, multiple GWAS studies were aggregated and the mean of λ was computed. **b)** Quantile-quantile plots of GWAS SNP p-values for dsbSNPs as compared to the expected distribution, and to the distribution of control SNPs (ctrlSNPs and ctrlSNPs (200-500kb)) for thyroid disease, primary biliary cholangitis, squamous cell lung carcinoma and gastric cancer. The study accession GSCTXXX is shown. **c)** Same figure as Fig. 6c but untruncated.

Next, we looked to assess the associations between dsbSNPs and individual diseases. To ensure enough statistical power, we did not restrict our analysis to GWAS top associations, but instead extracted all significant SNPs from summary statistics for all available disease-related studies in the GWAS Catalog. We compared the number of dsbSNPs significantly associated with GWAS-significant SNPs (*i.e.,* SNPs with p<10^−8^) to the other sets of SNPs using Fisher’s exact test (Fig. 6b; Sup. Table 1). Strikingly, we found 27/125 diseases for which the proportion of dsbSNPs mapping GWAS SNPs was significantly higher than for control SNPs (at FDR=5%). These diseases included several autoimmune diseases, such as thyroid disease, with 0.85% of significant dsbSNPs vs 0.23% of ctrlSNPs (OR=3.56, p=3.42×10^−7^) and 0.36% of ctrlSNPs (200-500kb) (OR=2.4, p=6.7×10^−5^), and primary biliary cirrhosis, with 0.84% of significant dsbSNPs vs 0.21% of ctrlSNPs (OR=3.66, p=6,5×10^−4^) and 0.31% of ctrlSNPs (200-500kb) (OR=2.61, p=0.0055). We also found cancers, including gastric carcinoma (vs ctrlSNPs: OR=8.41, p=1.45×10^−5^) and squamous cell lung carcinoma (SCLC) (vs ctrlSNPs: OR= 7.31, p=0.0016). In addition, we found that dsbSNPs were slightly more associated with diseases than eSNPs (0.0902% vs 0.0801%, p=0.03).

To confirm these associations, we then computed the genomic control factor (λ) for dsbSNPs and compared it to control SNPs and eSNPs (Fig. S6a). Overall, the λ was significantly higher than 1 (median=1.15, p<1×10^−16^), but not significantly higher than for eSNPs (median=1.11, p=0.1365). Among the previous diseases, thyroid disease, primary biliary cirrhosis, and squamous cell lung carcinoma showed a λ for dsbSNPs > 1.16, higher than for control SNPs and eSNPs (Fig. S6a). In agreement, qq-plots showed strong deviations from the expected distribution (Fig. S6b). In addition, we investigated the potential correlation between dsbSNP ES and GWAS ES and found that, depending on the disease, DSB-increasing alleles could be associated either with increased or decreased susceptibility (Sup. Info.).

We then assessed whether dsbSNPs were specifically associated with certain disease classes, using the GWAS Catalog disease classification (Fig. 6d). We only kept disease classes with 20 or more lambda values. As shown above, immune diseases and cancers showed a lambda average significantly higher than 1 (p=1.11×10^−7^, p=1.70×10^−9^, resp.) (Fig. 6c and Fig. S6c with the truncated and untruncated y-axes resp.). Strikingly, the lambda average was significantly higher than 1 for all the other disease classes, including respiratory diseases and cardiovascular diseases (all adjusted p-values < 0.05). Similar results were found for eSNPs.

Overall, these results showed a strong association of dsbSNPs with complex genetic diseases, and even stronger than for eSNPs. Although we initially hypothesized that dsbSNPs would be particularly associated with diseases linked to genome instability, we could show that dsbSNPs are associated with all classes of complex disease, suggesting a more central role of dsbSNPs in complex disease etiology.

## Conclusion

In summary, we mapped at high resolution more than 7,000 novel non-coding variants associated with DSB frequency (dsbSNPs), representing a significant fraction of variants in the genome, mainly found in promoters and introns. Using DNA motif analysis, we have investigated different causal mechanisms and found that dsbSNPs that regulate transcriptional activity affect the repair pathway choice. Given that NHEJ is more prone to errors than HR^54^, a SNP favoring NHEJ would tend to have a higher impact on the local mutation rate. In addition, we assessed the link between dsbSNPs and complex diseases and uncovered numerous disease associations overlapping dsbSNPs.

Here, we propose a novel mechanism by which non-coding SNPs contribute to complex genetic diseases by directly regulating genome instability and repair. Previous work, which had mapped quantitative trait loci associated with replication timing, known to be associated with genome instability, could not link them to DSB formation and explore the potential associations with complex diseases. This was likely due to the very low resolution of replication timing domains (100 kb)^23,24^, compared to our method which successfully mapped dsbSNPs at a resolution of ∼250bp. In addition, we found a two-fold enrichment of dsbSNPs at GWAS top associations compared to eSNPs, suggesting an important role of dsbSNPs in complex genetic diseases. This is especially relevant since Mostavafi et al. recently pointed out the surprisingly low overlap between eSNPs and GWAS hits and, using comprehensive statistical analyses, argued that a “fundamental issue is that GWAS and eQTL mapping are powered to identify different types of variants”^20^.

Previous works suggested that transcriptional activity could lead to DSB formation, potentially through transcriptionally-induced R-loop structures^50,56–58^. Using DNA motif-based causal analysis powered by our newly mapped dsbSNPs, we could not detect any causal effect of R-loops and transcription on DSB generation. Surprisingly, we instead uncovered that a dsbSNP disrupting a TF motif (and hence transcriptional activity) affects the choice of the repair pathway, in agreement with previous work showing a role of transcriptional activity in HR targeting at DSBs^53,59^. Besides transcription, DSB formation was previously associated with an increased TAD border fragility^46,47^. Using motif analysis, we found a causal yet weak effect of CTCF motif disruption on DSB formation, strengthening the previously proposed causal effect of the 3D genome on DSBs.

In this study, we have mapped several thousands of dsbSNPs. However, we are far from the amount of data generated for eSNPs by the GTEx consortium^14^. In particular, we have mapped dsbSNPs mainly in one cell line (U2OS-DIvA). Despite this limitation, we could already detect 1.3% of SNPs tested as dsbSNPs, and could demonstrate a strong link with complex diseases. Moreover, there was a significant overlap between dsbSNPs identified in GM12878 cells and dsbSNPs from U2OS-DIvA cells, and deep learning showed good generalization of DSB predictions across cell lines, further mitigating this limitation. Therefore, we speculate that dsbSNPs might be as pervasive as eSNPs in the genome. In this context, given the overlap between eSNPs and dsbSNPs, extensive studies are needed to better understand their interplay. In conclusion, such a study represents a proof of concept of the dsbSNP hypothesis, which will need further investigation in different disease contexts. We advocate for future studies to generate as much data as possible to map dsbSNPs in different normal tissues, cell lines, cancer, and patient samples.

## Materials and Methods

### Cell culture and treatments

We used U2OS-DIvA cells (modified U2OS osteosarcoma, [IacovoniJason+al:2010]), which were grown in Dulbecco’s modified Eagle’s medium (DMEM) supplemented with 10% SVF (Invitrogen), antibiotics, and 1 μg/ml puromycin at 37°C under a humidified atmosphere with 5% CO2. Cells were regularly tested and found negative for mycoplasma contamination.

### ChIP-seq data

In the U2OS-DIvA cell line, we have compiled a large database of ChIP-seq data from DNA repair proteins (pATM, 53BP1, RPA, SETX and Lig4), other proteins (CTCF, RAD21, SA2, RNAPII, RNAPII Ser2P, RNAPII Ser5P, RNAPII Ser7P, RNAPII T1P) and histone modifications (H3K27Ac, H3K4me1, H3K4me2, H3K4me3, H3K9me3, H4K20me2, H4K20me3, H2BK120ac, H2AZac) (Expression Array E-MTAB-5817, E-MTAB-8851, E-MTAB-6318) (Collins et al., in prep: E-MTAB-15639). Reads were aligned with BWA v0.7.17 on the human genome hg19 assembly^60^. Then BAM files were sorted, duplicates were removed, and files were indexed using samtools^61^.

We also downloaded ChIP-seq data from other cell lines: BRCA1 (GEO Accession: GSM935377), RAD51 (GEO Accession: GSE105699), and NBN (GEO Accession: GSE91629) in GM12878, and BRCA2 in hTert-HME1 (GEO Accession: GSE133450).

### ATAC-seq data

ATAC-seq experiments in the U2OS-DIvA cell line allowed us to identify chromatin accessible regions in this cell line (Collins et al., in prep: E-MTAB-15637). Reads were aligned with BWA v0.7.17 on the human genome hg19 assembly.

### DRIP-seq data

DRIP-seq experiments in the U2OS-DIvA cell line allowed us to identify R-loops in this cell line (Expression Array E-MTAB-11592). Reads were aligned with BWA v0.7.17 on the human genome hg19 assembly.

### DSB data

We also downloaded DSB data already mapped on hg19 from other cell lines: DSBCapture in NHEK cells (GEO Accession: GSE78172), sBLISS in neuronal cells (BioProject Accession: PRJNA798046), BLESS in untreated U2OS-DIvA cells (Expression Array E-MTAB-5817).

### DSB mapping

Given the large number of NGS methods to map DSBs, here we have recapitulated the different methods in the following table 1.

**Table 1:** Summary of the different methods to map DNA double-strand breaks (DSBs).

| DSB mapping | Based on ChIP-seq | Reference |
| --- | --- | --- |
| DNA repair proteins: pATM, 53BP1, SETX, RPA, Lig4, RAD51, NBN, BRCA1, BRCA2. | Yes | 62 |
| DSBCapture | No | 35 |
| BLESS | No | 1 |
| sBLISS | No | 37 |
| END-seq | No | 3 |

### Heterozygous SNP calling

We used the ADASTRA pipeline^16^. Briefly, BAM alignment files from which duplicates have been removed with Picard were supplied to GATK 4.2.1.0 for base quality score recalibration followed by GATK HaplotypeCaller with default parameters (https://gatk.broadinstitute.org/hc/en-us). The resulting variant calls were filtered according to the following criteria: (1) heterozygous variants (GATK GT=0/1), (2) with coverage of more than five reads supporting the reference and five supporting the alternative alleles, and (3) found in dbSNP v.151 (GRCh37.p13 genome assembly) common variants subset (https://www.ncbi.nlm.nih.gov/snp/). In order to account for potential copy number variations (CNVs), we have constructed genomic maps of the background allelic dosage (BAD) as in ADASTRA with BABACHI 2.0^16^ solely from ChIP-seq data.

### Detection of allele-specific SNPs

Identification of the allele-specific variants from allelic imbalance consists of detecting a significant difference in NGS (e.g., ChIP-seq) read counts between the two alleles (e.g., the reference and alternative alleles) at heterozygous loci of homologous chromosomes in a single diploid genome.

An allele-specific SNP is determined through comparison of the number of reads supporting the reference (Nref) and the alternative (Nalt) alleles. For a heterozygous SNP, assuming no CNV, the expected allelic ratio 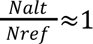, and a significant deviation from 1 corresponds to the allelic imbalance. In the case of a CNV, the expected ratio may be different than 1, and allelic imbalance is estimated against the expected ratio corrected for the CNV.

To statistically assess the allelic imbalance at each SNP, we used MIXALIME version 2.15.1^63^ with the default parameters, except default-bad=6 to penalize SNPs located outside of the regions included in the BAD map. For scoring SNPs, we used a mixture of negative binomial (for ChIP-seq) or beta negative binomial (for ATAC-seq) distributions. The resulting beta negative binomial mixture model for ATAC-seq was more conservative as it accounted for the extra overdispersion. DRIP-seq data were scored with the same model as ChIP-seq. The candidate SNPs for further analysis were filtered by requiring a total coverage of 20 reads in at least one sample, to reduce false positive SNP calls. Next, SNP-level p-values of the allelic imbalance were combined across different samples for Ref- and Alt-alleles separately using the Mudholkar-George method^64^ and FDR-adjusted for multiple tested SNPs by the Benjamini-Hochberg procedure^65^. This was performed for each data type separately. We used a threshold of 10% for the FDR to select the significant allele-specific SNPs. Finally, the ADASTRA-style estimates for Ref- and Alt-skewed allele-specific variants were merged at each particular SNP by selecting the preferred allele with a lower p-value.

For each SNP, we used individual estimates of effect size for each ChIP-seq experiment as provided by MIXALIME. When there were multiple ChIP-seq experiments for a single protein, for each SNP for the effect size estimate we used the ES with the maximum absolute value across replicates.

### ADASTRA database of allele-specific protein binding

In this study, we used the ADASTRA database v5.1.3 on allele-specific protein binding^16^ for several proteins of interest, namely ATM, BRCA1, BRCA2, NBN and RAD51.

### SNP filtering

We filtered out the SNPs at centrosomes and telomeres. We also removed SNPs that were too close to an AsiSI site of the U2OS-DIvA cell line (in order to minimize potential bias resulting from basal enzyme activity, even without treatment). For pATM SNPs, we filtered out those that were less than 1kb from an AsiSI site. For Lig4 SNPs, we filtered out those that were less than 20kb from an AsiSI site. For 53BP1, we filtered out those that were from less than 500kb from an AsiSI site.

### Genome browser

To browse genomic regions, we used Integrative Genomics Viewer (IGV) (https://igv.org/). To display the number of reads for each allele of a dsbSNP, we proceeded as follows. We first counted the number of reads containing each allele. More importantly, to account for potential copy number variations (CNVs), we then divided the number of reads of the alt allele by the expected ratio 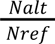, as computed previously in subsection Detection of allelic-specific SNPs.

### SNP correlation heatmap

We calculated the Pearson correlation coefficient of SNP effect size for each pair of ChIP-seq experiments (DNA repair binding proteins, TF proteins, and histone marks). Correlations with less than 20 data points (ie, SNPs) were set to NA.

### Annotation of SNPs

For the annotation of SNPs, we used the R function annotatePeak() from the ChIPseeker package^66^.

### Average profiles

Deeptools was used to compute average profiles of read coverage over sets of SNPs (e.g., dsbSNPs)^67^.

### GTEx SNPs (normal tissues)

We downloaded cis-expression SNPs (cis-eSNPs) identified from different tissues from the Genotype-Tissue Expression (GTEx) v7 project (https://www.gtexportal.org/home/index.html)^14^.

### PancanQTL SNPs (cancers)

We downloaded cis-expression SNPs (cis-eSNPs) identified in various cancers from The Cancer Genome Atlas (TCGA) project (http://gong_lab.hzau.edu.cn/PancanQTL/)^68^.

### GWAS summary statistics

Summary statistics of SNPs associated with common genetic diseases identified by genome-wide association studies (GWASs) were downloaded from NHGRI-EBI GWAS catalog on 12 July 2022 (https://www.ebi.ac.uk/gwas/)^55^. GWAS disease classes were also downloaded.

### Control SNPs

For comparison purposes, we used three types of control SNPs from the 1000 genomes project with a MAF>10%, either randomly picked in the genome (“ctrlSNPs”), or randomly picked with less than 500 kb from dsbSNPs (“ctrlSNPs (<500kb)”), or randomly picked within a 200-500 kb distance from dsbSNPs (“ctrlSNPs (200-500kb)”).

### Q-Q plot

The Q-Q plot was used to graphically show the association of dsbSNPs with a given GWAS study. For this purpose, the GWAS SNP p-values for the dsbSNPs were extracted and compared with the expected distribution or the GWAS SNP p-values for the control SNPs.

### Association with GWAS diseases

We tested the association between dsbSNPs and GWAS hits for each disease using summary statistics. For this purpose, for a given disease, we first extracted all p-values from summary statistics. To secure enough statistical power, we only kept studies with enough (500) SNPs with p < 10^−6^, which resulted in 227 studies. We then counted the number of dsbSNPs with a p-value < 10^−8^ versus the number of control SNPs with a p-value < 10^−8^ (as compared to the number of dsbSNPs with a p-value > 10^−8^ and the number of control SNPs with a p-value > 10^−8^). Since there could be multiple GWAS studies for a given disease, we aggregated the counts and also added a pseudocount of 1 to compute the odds ratio (OR). To assess significance, we used the unilateral Fisher’s exact test (without pseudocount), and computed adjusted p-values by false discovery rate (FDR).

### Genomic control (λ)

The lambda for genomic control was computed as the median of p-values divided by 0.456 using R base.

### Transcription factor binding DNA motifs

Known protein-binding DNA motifs from JASPAR 2024 database (https://jaspar.elixir.no/) were scanned at dsbSNP positions with a context of 101 bp (+/− 50 bp around the dsbSNP) using the R package motifmatchr (https://bioconductor.org/packages/release/bioc/html/motifmatchr.html). We used the set of non-redundant motifs.

For both alleles of a dsbSNP, positive weight matrix scores were computed for a given protein-binding DNA motif. For each allele, a window of 25 bp was used (+/− 12 bp around the dsbSNP). Only motifs that overlap the SNP in the 25 bp window were kept. The motif delta score was computed as the difference between the score of the alternative allele and the score of the reference allele.

### Non-B DNA motifs

We downloaded non-B DNA motifs mapped on hg19 from https://nonb-abcc.ncifcrf.gov. We computed GC-skew with a window of 501 bp (+/− 250 bp around the dsbSNP) as : (G - C)/(G + C).

### Deep learning

We used Selene to train deep learning DeepSea models to predict ChIP-seq and DSBCapture peaks from DNA sequences (https://github.com/FunctionLab/selene/)^33^. Here, we used DeeperDeepSea models with only one classification head. We kept the default parameters, including using chromosomes 8 and 9 for the test held-out dataset. Using the trained models, we predicted the effect of SNPs on the presence of a peak.

## Data availability

All high-throughput sequencing data generated (ChIP-seq, DRIP-seq, and ATAC-seq) have been deposited to Array Express (https://www.ebi.ac.uk/arrayexpress/) under accession E-MTAB-15639, E-MTAB-15637.

## Code availability

The source code is available on GitHub:https://github.com/raphaelmourad/dsbSNP.

