## Supplementary material for "A novel class of non-coding variants driving DNA double-strand breaks is associated with complex genetic diseases": Supp Info 1

### Supplementary Information

#### Correlation between GWAS effect sizes and dsbSNP effect sizes

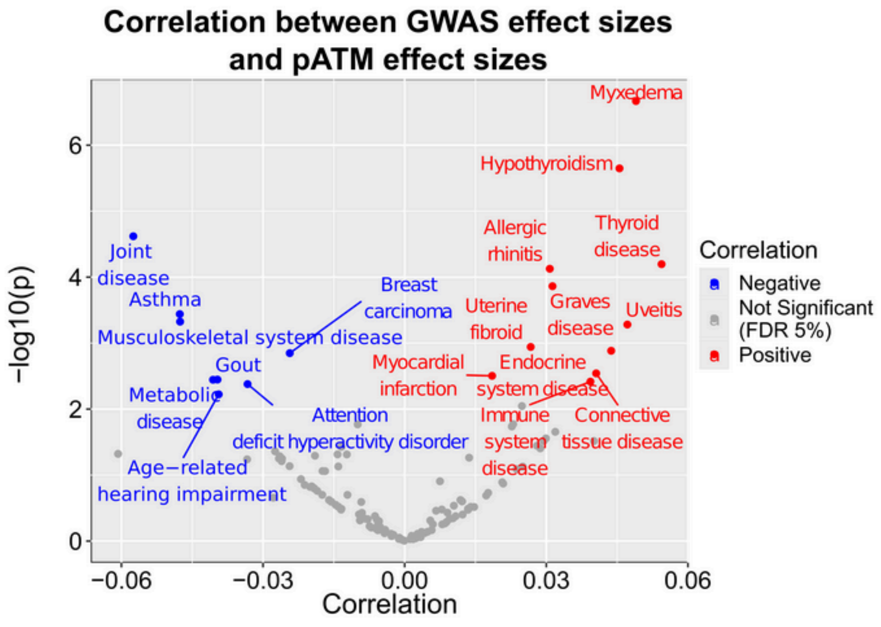


Figure SI1: Volcano plot showing the correlation between pATM SNP effect size and GWAS SNP effect size, and corresponding p-value.

We investigated a potential positive or negative effect of DSB-increasing alleles with disease susceptibility. For this purpose, we assessed the Pearson correlation between pATM SNP effect sizes and GWAS SNP effect sizes for each disease. Overall, there was no clear trend for the correlations ($H_0$: r=0, p=0.97). However, interestingly, for myxedema, we found a positive correlation (r=0.049, p=2x$10^{-7}$, corrected p=3x$10^{-5}$; Fig. SI1), while for joint disease and asthma, we found negative correlations (r=-0.057, p=2x$10^{-5}$, corrected p=0.001; r=-0.048, p=4x$10^{-4}$, corrected p=0.007, resp.; Fig. SI1). Such results suggest that, depending on the disease, DSB-increasing alleles could be associated either with increased or decreased susceptibility.
